# Rapid classification of a novel ALS-causing I149S variant in superoxide dismutase-1

**DOI:** 10.1101/2024.01.17.23300408

**Authors:** VK Shephard, ML Brown, BA Thompson, A Harpur, L McAlary

**Affiliations:** Molecular Horizons and School of Chemistry and Molecular Bioscience, Faculty of Science Medicine and Health, University of Wollongong, NSW 2522, Australia; Department of Pathology, Royal Melbourne Hospital, Melbourne, VIC 3050, Australia; Department of Genomic Medicine, Royal Melbourne Hospital, Melbourne, VIC 3050, Australia

## Abstract

Variants of the oxygen free radical scavenging enzyme superoxide dismutase-1 (SOD1) are associated with the neurodegenerative disease amyotrophic lateral sclerosis (ALS). These variants occur in roughly 20% of familial ALS cases, and 1% of sporadic ALS cases. Here, we identified a novel SOD1 variant in a patient in their 50s who presented with movement deficiencies and neuropsychiatric features. The variant was heterozygous and resulted in the isoleucine at position 149 being substituted with a serine (I149S). *In silico* analysis predicted the variant to be destabilising to the SOD1 protein structure. Expression of the SOD1^I149S^ variant with a C-terminal EGFP tag in neuronal-like NSC-34 cells resulted in extensive inclusion formation and reduced cell viability. Immunoblotting revealed that the intramolecular disulphide between Cys57 and Cys146 was fully reduced for SOD1^I149S^. Furthermore, SOD1^I149S^ was highly susceptible to proteolytic digestion, suggesting a large degree of instability to the protein fold. Finally, fluorescence correlation spectroscopy and native-PAGE of cell lysates showed that SOD1^I149S^ was monomeric in solution in comparison to the dimeric SOD1^WT^. This experimental data was obtained within 3 months and resulted in the rapid re-classification of the variant from a variant of unknown significance to a clinically actionable likely pathogenic variant.

## Introduction

Amyotrophic lateral sclerosis (ALS) is a rapidly progressive neuromuscular disease in which motor neurons within the upper and lower spinal cord degenerate^1^. The loss of motor neurons leads to weakness, spasticity, and eventually paralysis, often resulting in patient death roughly 2.5 years after diagnosis ^2^. The majority of ALS cases are sporadic (90%), however, some are associated with a familial history of disease (FALS; 10% of cases). Of the FALS cases, around 20% are associated with amino acid variants in the cytosolic oxygen free radical scavenger protein superoxide dismutase-1 (SOD1) ^3^. There are now over 220 SOD1 variants listed to be potentially ALS-associated in the ALS online database, with the majority of these variants being single site substitutions spread throughout the protein sequence^4^. The major hypothesis for how variants of SOD1 cause ALS is that variants destabilise SOD1 folding stability, resulting in misfolding and aberrant protein aggregation^5,6^. This is supported by the existence of insoluble SOD1 inclusions in the post-mortem tissue of patients who suffered from SOD1-associated FALS^7^.

Some SOD1 variants exist in populations without causing ALS whilst others are known to invariably cause ALS^8^. A difficulty in the clinical landscape of SOD1-associated ALS has been acquiring evidence to accurately classify rare/novel variants of SOD1 in ALS patients as pathogenic on the basis of clinical evidence alone^9^. Therefore, molecular and biochemical analyses need to be utilised to aid in the classification of rare/novel variants to enhance outcomes for patients. Rapid and accurate classification allows patients to be more clearly diagnosed and more easily access appropriate care^10^. Additionally, the most promising current ALS treatment is an antisense oligonucleotide targeted against SOD1 (toferson)^11^, meaning that if a patient can be identified they may receive treatment quicker. Furthermore, some clinical trials are aiming to treat pre-symptomatic patients with toferson (NCT04856982), in which knowledge about more SOD1 variants is essential in expanding trial coverage and potentially affording greater efficacy. Therefore, methods to quickly and accurately classify SOD1 variants are of high importance to facilitating understanding and treatment of ALS.

Here, a patient in their 50s presented with clinically manifest neurological and movement deficiencies. Specifically, the patient suffered from a chronic gradually progressive form of a neurodegenerative disorder characterised by choreiform movement, anterior horn cell changes, frontal lobe cognitive changes, neuropsychiatric features, right sided weakness, unsteady gait. Two years after initial presentation, genetic sequencing identified a novel SOD1 variant that resulted in the isoleucine at position 149 mutating to serine (NM_000454.4:c.449T>G p.(Ile149Ser)) in the primary sequence, henceforth called SOD1^I149S^. Molecular and biochemical analysis of SOD1^I149S^ showed that the variant readily formed inclusions in NSC-34 cells, similar to other known SOD1-FALS variants. NSC-34 cells expressing SOD1^I149S^ had significantly reduced viability compared to cells expressing wild-type SOD1 (SOD1^WT^) Furthermore, our biochemical analysis showed that SOD1^I149S^ did not form the intramolecular disulphide, did not form dimers, and was substantially destabilised in comparison to SOD1^WT^. Collectively, these data were used to reclassify the variant from uncertain significance to likely pathogenic in under 3 months from identification of the variant via sequencing.

## Results

### Patient Clinical Examination

The patient presented with a complex clinical picture, including neurological symptoms such as choreiform movements, anterior horn cell changes, cognitive alterations, and neuropsychiatric features. Initially, the patient reported significant weight loss, slurred speech, difficulty swallowing, muscle twitching, cramps, and marked right-sided limb weakness, resulting in multiple falls. Mood disturbances, disorganization, impulse control issues, poor judgment, and chronic sleep problems were also observed.

On examination, the patient displayed choreiform movements, delayed saccades, dysarthric speech, fasciculations, and muscle wastage in the right shoulder. Gait abnormalities, including a narrow-based gait with high-stepping pattern, were evident, while no dystonia or parkinsonism was observed. Additional neurological exams were unremarkable.

Extensive blood tests and scans yielded no abnormalities or evidence of malignancy. Electromyography indicated fasciculations, and genetic testing for Huntington’s disease and *C9orf72* hexanucleotide repeat expansions returned negative. Genetic testing for *SOD1* identified a heterozygous variant in the *SOD1* gene (NM_000454.4:c.449T>G p.(Ile149Ser)) which was then further investigated by biochemical methods.

### SOD1^I149S^ is predicted to be destabilising and decrease aggregation

SOD1 monomers adopt a well-structured and highly stable eight-stranded greek key conformation when fully mature^5,6^. A crystal structure of SOD1 shows that I149 is oriented so that the sidechain is positioned within the hydrophobic core of the protein **(Figure 1A)**, suggesting that its change to a polar residue could potentially disturb residue I149’s contribution to the formation of the SOD1 hydrophobic core^12^. Additionally, SOD1 contains multiple amyloidogenic regions within its sequence, one of which is localised to the far C-terminus, incorporating sequence segment _147_GVIGIAQ_153_^13^. Input of the sequences for SOD1^I149S^ and SOD1^WT^ into the amyloid prediction tool Aggrescan 2D^14^ showed that the I149S substitution is predicted to reduce the amyloidogenicity of the C-terminus of SOD1 **(Figure 1B)**, but not completely.

**Figure 1.**
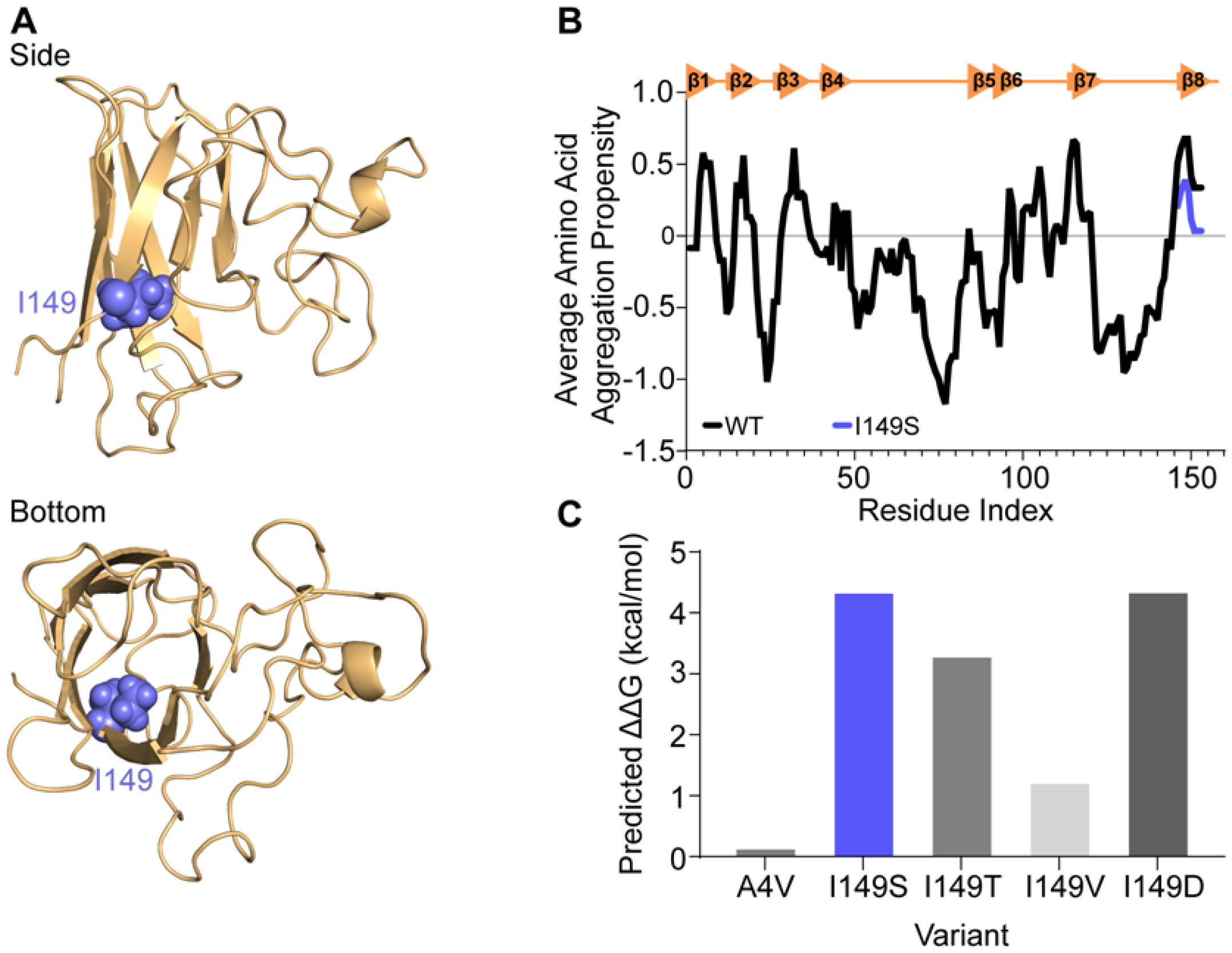
*In silico* analysis of the effect of amino acid substitution at position I149 of SOD1 predicts lower aggregation propensity but greater instability for a SOD1^149S^ variant. **(A)** Side and bottom views of the SOD1 monomer structure showing residue I149 as spheres (blue) oriented into the core of the beta-barrel. **(B)** Aggrescan2D measurement of wild-type (WT; black) and I149S (blue) on the aggregation propensity of SOD1. **(C)** MAESTRO predicted folding stability changes of the variants at position I149 in SOD1 and A4V, showing that amino acid subsitution at I149 is predicted to be destabilising.

Further modelling was performed using the MAESTRO web server^15^ for prediction of the effect of the SOD1^I149S^ variant on the stability of the protein. We also incorporated several variants into the predictions including known ALS-causing variants (SOD1^I149T^, SOD1^I149V^, and SOD1^A4V^) and a synthetic variant, SOD1^I149D^. MAESTRO predicted that SOD1^I149S^ was a highly destabilising variant, resulting in a predicted ΔΔG value of 4.35 kcal/mol **(Figure 1C)**. In comparison, SOD1^A4V^ is predicted by MAESTRO to destabilise the protein by only 0.15 kcal/mol, suggesting that SOD1^I149S^ may be potentially more pathogenic **(Figure 1C)**. Other disease-causing variants localised to I149 are SOD1^I149T^ and SOD1^I149V^. We tested these variants in MAESTRO as well, finding that each were predicted to be destabilising, with SOD1^I149T^ decreasing predicted stability by 3.30 kcal/mol and SOD1^I149V^ decreasing stability by 1.23 kcal/mo **(Figure 1C)**. We also designed a variant that would introduce charge into the SOD1 beta barrel core, I149D, which was predicted to destabilise the SOD1 fold with a ΔΔG value of 4.36 kcal/mol.

### SOD1^I149S^ forms inclusions and is toxic in a motor neuron-like cell model of SOD1-FALS

Previous reports have shown that when SOD1 variants are overexpressed in established cell lines, they can form insoluble inclusions similar to those found in patients post-mortem^16^. Indeed, this cellular model is used routinely to investigate variants and the effect of small molecules on SOD1 inclusion formation and toxicity^17^. In order to determine if the SOD1^I149S^ variant led to inclusion formation, we transiently expressed it in NSC-34 cells with a C-terminal EGFP tag alongside other variants SOD1^WT^, SOD1^A4V^, SOD1^I149D^, SOD1^I149T^, and SOD1^I149V^. All of the variants are known FALS variants except for SOD1^I149D^ which is a synthetic variant designed to maximally destabilise the I149 site. Fluorescence microscopy and automated image analysis were used to determine the percentage of cells that contained inclusions and enumerate the number of GFP-positive cells across time to determine relative cell numbers^18^.

We measured inclusion formation in the top 10% highest expressing cells (determined by measuring GFP intensity per cell), as we have previously determined this population to be the most likely to form inclusions^19^. We observed that all variants formed significantly more inclusions than SOD1^WT^ (P < 0.001) after 48 h post-transfection **(Figure 2B)**. Expression of the novel variant SOD1^I149S^ resulted in approximately 30% of cells containing inclusions, which was higher than those cells expressing SOD1^A4V^, where approximately 23% of cells contained inclusions. The relative effect of the SOD1 variants on cell health was determined by enumerating the number of GFP-positive cells at 24 h, 48 h, and 72 h post-transfection and normalising the values for each variant to cell counts from the 24 h time point, and the SOD1^WT^ counts **(Figure 2 C)**. From this, we observed that variants SOD1^A4V^, SOD1^I149T^, SOD1^I149S^, and SOD1^I149D^ all resulted in a loss of GFP-positive cells over time, indicating that those cell populations had their growth affected by expression of these variants **(Figure 2C)**. SOD1^I149V^ did not differ substantially from SOD1^WT^ at any time point, indicating this variant was not affecting cell growth. Statistical analysis of the GFP-positive cells counts from the 72 h time point showed that SOD1^A4V^, SOD1^I149T^, SOD1^I149S^ and SOD1^I149D^ were significantly lower at this point when compared to SOD1^WT^ **(Figure 2D)**.

**Figure 2.**
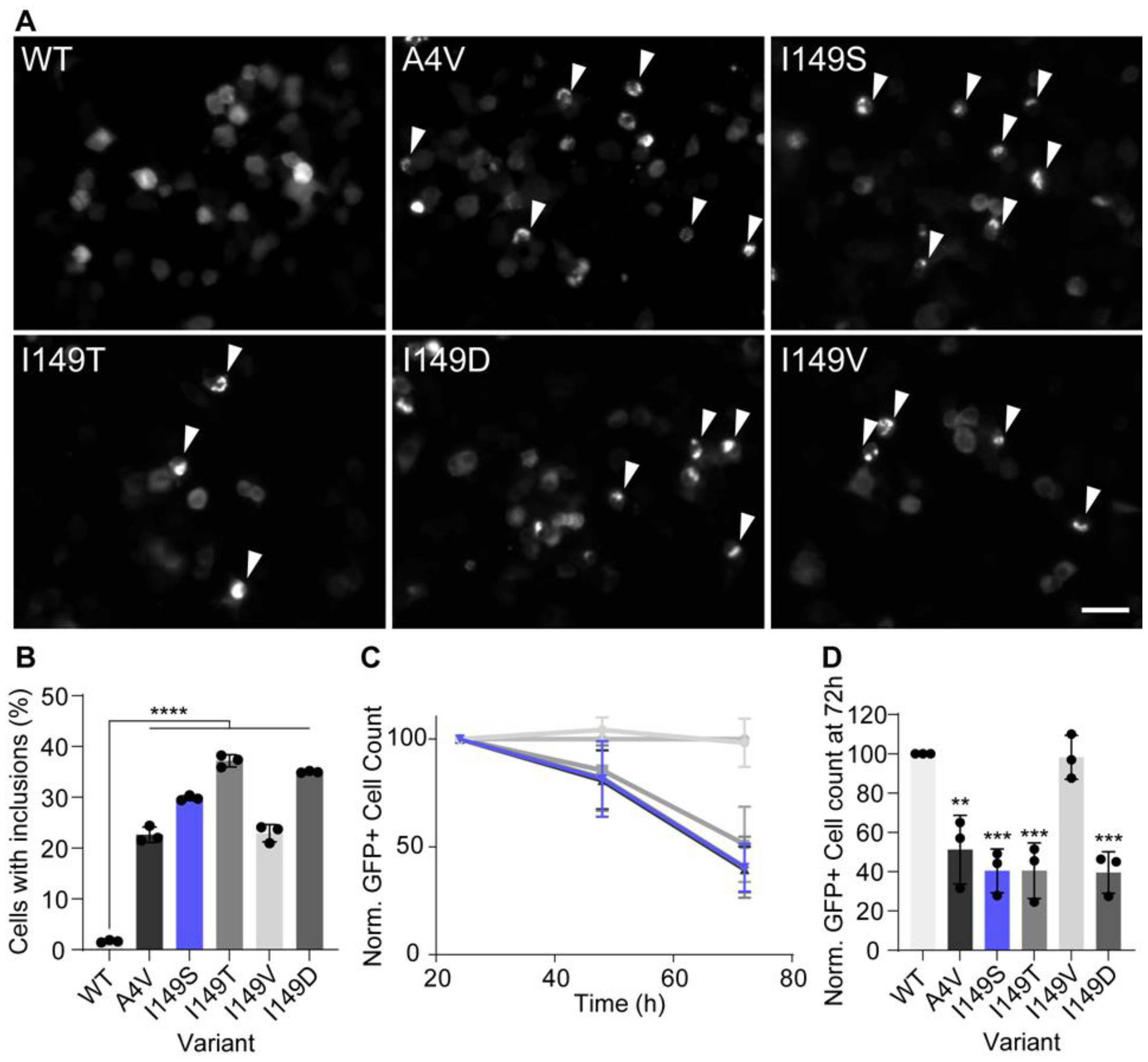
SOD1^I149S^ forms inclusions and decreases the viability of motor neuron-like cells. **(A)** Example images of NSC-34 cells expressing GFP-tagged SOD1 variants. Arrows point to cells containing inclusions. Scale bar = 50 μm. **(B)** The percentage of cells containing inclusions within the top 10% highest expressing cells, showing the I149S readily forms inclusions. **(C)** Enumeration of GFP+ cells 24, 48, and 72 h post-transfection normalised to 24 h and wild-type counts, showing that cells expressing I149S (blue) decrease over time. **(D)** Quantification of the 72 h time point from panel C, showing that at 72 h, SOD1-I149S expressing cells are at 40% the number of cells expressing SOD1-WT. Error bars represent SD from at least 3 biological replicates. Significance determined by One-way ANOVA with Tukey’s post test. **** = P < 0.0001, *** = P < 0.001, and ** = P < 0.01.

### SOD1^I149S^ is highly destabilised when expressed in a cell model of SOD1-FALS

The primary consequence of variants in SOD1 is the destabilisation of the protein fold through various interconnected biophysical features of the protein^5,6^. These features include the general folding stability of the protein, the intramolecular disulphide bond, and dimerisation. All of these features can be measured biochemically to determine how a variant may be affecting the folding state of SOD1 in cells.

To examine the consequence of the variants on SOD1 disulphide formation, we exploited the differential electrophoretic mobility of disulphide bonded SOD1 in comparison to reduced SOD1 **(Figure 3 A and B)**^20^. We found that both SOD1^WT^ and SOD1^I149V^ had similar levels of disulphide formation at approximately 40% of measured SOD1. Meanwhile, SOD1^A4V^ showed significantly lower (P < 0.0001), but still measurable, levels of disulphide formation at approximately 10% of protein. Only around 2% of SOD1^I149T^ was found to be disulphide bonded, indicating severe alteration to the ability of SOD1^I149T^ to undergo or maintain this post-translational modification. Both SOD1^I149S^ and SOD1^I149D^ had virtually no detectable levels of disulphide bonded SOD1, further indicating a substantial effect of these variants on this important post-translational modification. Statistical analysis showed that all variants except SOD1^I149V^ had significantly less disulphide bonded SOD1 present (P < 0.0001).

**Figure 3.**
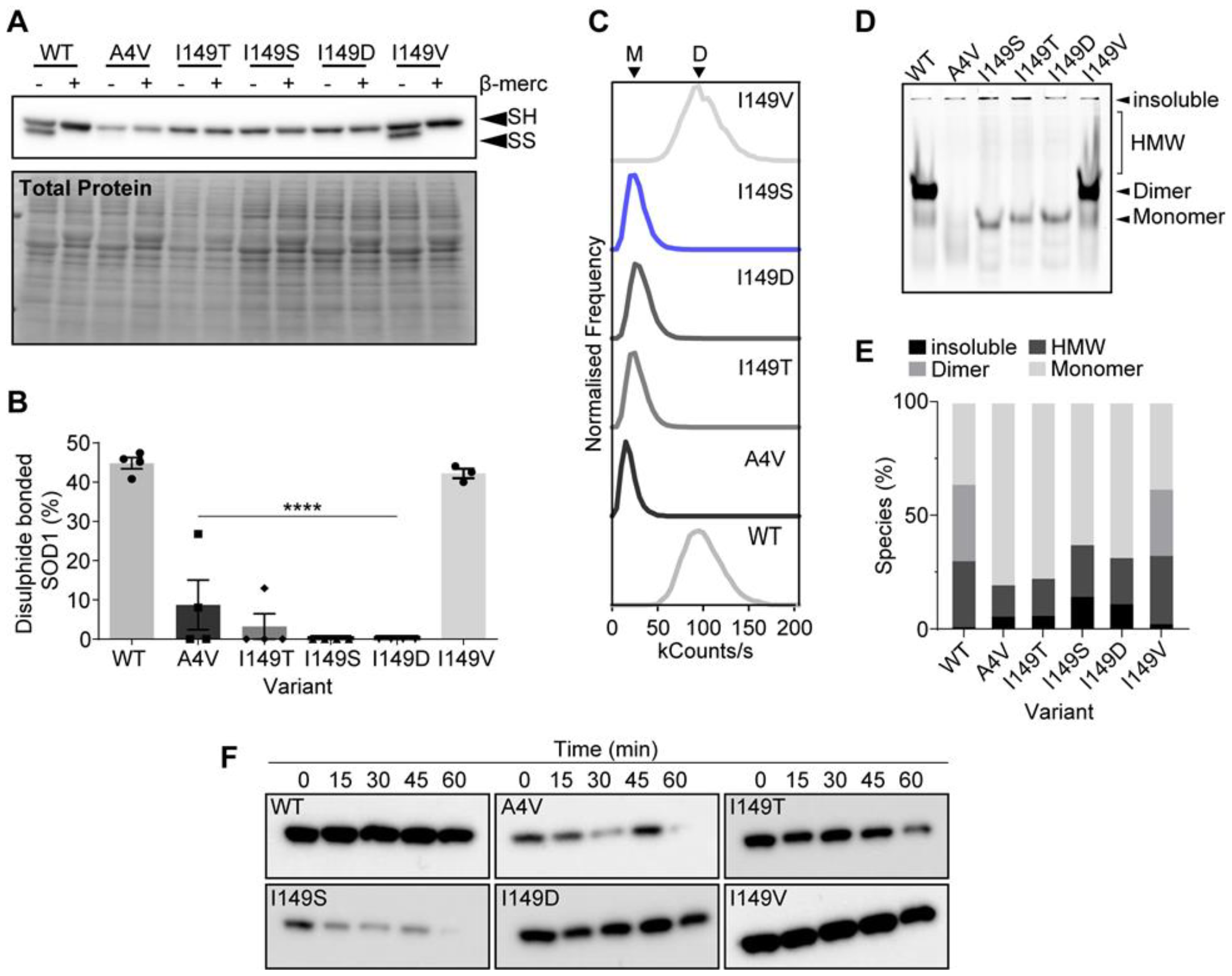
SOD1^I149S^ destabilises the intramolecular disulphide and decreases dimer formation. **(A)** Immunoblotting for disulphide bonded SOD1 was carried out by running non-reducing SDS-PAGE, where faster migrating bands (SS) are disulphide bonded SOD1 and slower migrating bands (SH) are disulphide reduced SOD1. **(B)** Quantification of the percentage of SOD1 that was diulphide-bonded showing that SOD1^I149S^ was completely reduced. **(C)** Brightness histograms from fluorescence correlation spectroscopy showing that the average brightness of SOD1^WT^ and SOD1^I149V^ were higher than other variants, indicating these two variants are oligomeric. **(D)** Clear Native-PAGE of transfected cell lysates showing GFP signal where several species are observable (HMW = high molecular weight). Only SOD1^WT^ and SOD1^I149V^ showed any dimer signal. **(E)** Quantification of the percentage of each species present from clear Native-PAGE in panel (D). **(F)** Limited proteolysis of SOD1 variants using proteinase-K showed that the variants were differentially susceptible to proteolysis over 1 h. Error bars represent SD of the mean from 3 separate experiments. Significance determined with One-way ANOVA using Tukey’s post-hoc test (P < 0.0001 = ****).

We next examined the effect of several variants on the ability of SOD1 to form native dimers. To do this, we utilised fluorescence correlation spectroscopy (FCS) and clear Native-PAGE. FCS is a technique that can determine the molecular brightness of molecules in solution^21^. In our case, a dimer of SOD1-EGFP would be expected to be brighter than a monomer. We observed that both SOD1^WT^ and SOD1^I149V^ had a molecular brightness of roughly 100 kcounts/s, whereas the other variants all were found to have molecular brightness readings of around 25 kcounts/s **(Figure 3 C)**. The lower count rate for some variants indicates that the detected molecules produced less photons, suggesting fewer GFP molecules (monomers) were being detected for a single event. In conjunction, clear Native-PAGE separated SOD1-EGFP variants on their charge and hydrodynamic volume, where both SOD1^WT^ and SOD1^I149V^ showed an intense band that represents dimer **(Figure 3 D and E)**. Other variants did not show this band, but rather only monomer bands, high molecular weight smears, and signal in the gel wells that is insoluble material too large to enter the gel matrix. Together, FCS and clear Native-PAGE indicate that soluble SOD1^I149S^ is monomeric in cell lysates.

Finally, we examined the susceptibility of these SOD1 variants to proteolytic digestion from proteinase-K. Past research has shown that SOD1^WT^ is resistant to proteolysis whereas ALS-associated variants are susceptible to proteolysis due to the structural destabilisation induced by amino acid substitutions to the protein^22^. Lysates from cells expressing SOD1 variants were harvested and incubated for different times prior to immunoblotting for detection of SOD1-EGFP. We found that SOD1^WT^ and SOD1^I149V^ were both resistant to proteolytic digestion via proteinase-K. SOD1^I149D^ was only partially susceptible to digestion, whereas SOD1^A4V^, SOD1^I149T^, and SOD1^I149S^ were all susceptible to proteolytic digestion, indicating these variants are highly structurally destabilised. Collectively, this data was used to reclassify the variant to being likely pathogenic.

## Discussion

In this work we identified a novel SOD1 variant I149S, which we compared in cell and biochemical assays to several other variants, including those occurring at the same amino acid position. I149 is located on beta-strand 8 of the SOD1 protein, which takes part in the formation of the SOD1 dimer interface and is proximal to C146, which is responsible for intramolecular disulphide bond formation^5,6^. Consequently, we measured that the SOD1^I149S^ variant substantially altered these two features of SOD1 structure compared to SOD1^WT^. Impaired dimerization^23^ and presence of reduced SOD1^24^ are hallmarks of other pathological variants of SOD1. Indeed, SOD1^I149S^ was found to be even more destabilising than the prevalent and rapidly progressive SOD1^A4V^ variant in our assays. Curiously, SOD1^I149V^ behaved similarly to SOD1^WT^ in all assays except the inclusion formation assay. Our expectation was that SOD1^I149V^ would be less destabilising due to the amino acid substitution maintaining a hydrophobic residue. We did not expect such extensive inclusion formation without other features of the protein being affected. Previous research has suggested that the presence of inclusions does not always correlate with disease, and our analysis of SOD1^I149V^ supports this^25^. Overall, our assays were capable of capturing the diverse outcomes that occur with even just different residue substitutions at the same site in SOD1.

Using our robust and comprehensive assays, in under 3 months, we were able to effectively reclassify SOD1^I149S^ as ‘likely pathogenic’ according to ACMG/AMP criteria^9^. As a result, the patient now has a clear diagnosis, an understanding on his prognosis, potential access to clinical trials, and his relatives now have the choice of predictive testing. Currently, around 50% of known SOD1 variants are listed as only having *in silico* predictions of their pathogenicity, evidence that is not recommended to classify a variant alone. In the absence of appropriate clinical evidence, functional assay evidence is possibly the only other evidence available to classify a variant of uncertain significance. It has been three decades since variants in SOD1 were the first identified genetic linkage to ALS^26^. Since then, over 220 SOD1 variants have been linked to ALS with varying degrees of evidence to support their pathogenicity. Roughly 7 new variants are identified every year world-wide, yet the evidence needed to accurately classify these variants according to standard ACMG/AMP criteria is routinely lacking. Often, variants are identified in patients but not characterised for many years, or at all, due to poor communication between clinicians and basic scientists, and a lack of effective and rapid models for variant phenotype assessment^27^.

Considering the most promising therapies in clinical trials for ALS are currently targeted at SOD1^11^, increasing the entry of patients into these clinical trials is highly important to enhance trial outcomes. A clinical trial that has recently started in Australia is the ATLAS trial, which aims to use the SOD1-targeting antisense therapy, toferson, on pre-symptomatic SOD1 variant carrying patients (NCT04856982). Classification of the current SOD1 VUS pool would enhance the entry of patients into such clinical trials. Furthermore, prior knowledge and accurate classification of SOD1 variants is essential if we are to begin treating SOD1-FALS in people prior to symptom onset, which is the likely best therapeutic avenue available for ALS. In this respect, the methods we used here are cheap and effective for this purpose. In regards to SOD1 and other diseases, we propose that clinicians and basic researchers should more readily work together to solve patient-specific problems with diagnosis and treatment moving forward. This would greatly improve outcomes for patients and enhance the body of knowledge surrounding diseases such as ALS going forward.

## Methods and Materials

### Genetic Analysis

Exome sequencing was performed by Australian Genome Research Facility Ltd (AGRF) with the SureSelect XT Low Input Clinical Research Exome V2 (Agilent Technologies) kit and sequencing on an Illumina NovaSeq6000, with a targeted mean coverage of 100x and a minimum of 95% of bases sequenced to at least 10x. Data were processed at AGRF using their implementation of the GATK (v4.0) best-practice pipeline including HaplotypeCaller, in order to generate annotated variant calls within the target region (coding exons), via alignment to the reference genome (GRCh38). The following phenotype-driven virtual gene panels from PanelApp Australia (https://panelapp.agha.umccr.org/panels/) were used to assess relevant variants: Early-onset Dementia v0.154, Motor Neurone Disease v0.126, and Myopathy Superpanel v1.82. Variant prioritization and reporting were performed using Alissa Interpret v5.4.2 (Agilent Technologies). Coding variants were annotated against all gene transcripts, with reporting of variants against the MANE Select v1.0 recommended transcript. Classification of variants is based on modified ACMG/AMP guidelines^9^.

### In Silico Analysis

The canonical SOD1 amino acid sequence, without an N-terminal methionine, was obtained from Uniprot (Accession No. P00441) for input into online *in silico* sequence analysis tools. Variants were generated by replacing amino acids for A4V, I149S, I149T, I149V, and I149D. Aggrescan 2D was used to determine the amyloid aggregation potential of the SOD1 variants in comparison to WT via the windowed averaged amino acid aggregation propensity (a4v) output. The MAESTRO server was used to predict stability changes induced by variants in SOD1 via input of the SOD1^WT^ and variant sequences. In MAESTRO a positive ΔΔG value indicates destabilisation of the protein fold.

### Plasmids

Vectors for the expression of C-terminally EGFP-tagged SOD1 variants SOD1^WT^, SOD1^A4V^, SOD1^I149D^, SOD1^I149S^, SOD1^I149V^, SOD1^I149T^ on a pEGFP-N1 backbone were generated as described previously ^16^. Plasmids were heat transformed into chemically competent *E. coli* DH5α cells and purified from bacterial culture using maxiprep kits (Thermo Fisher Scientific) as per manufacturer’s instructions.

### Maintenance and Transfection of Mammalian Cells

Neuroblastoma × spinal cord (NSC-34) cells were cultured in Dulbecco’s modified Eagle’s medium-F12 (DMEM-F12) (Invitrogen) supplemented with 10% (v/v) heat inactivated foetal bovine serum (Bovagen). NSC-34 cells were passaged using 0.25 % trypsin and 0.02 % EDTA dissociation reagent (Invitrogen) to lift cells, then pelleted via centrifugation for 5 minutes at 300 g. The cell pellet was resuspended in prewarmed culture media, seeded at 40 % confluency into a 6 well plate and grown at 37 °C in a humidified incubator with 5 % atmospheric CO_2_ for 24 h. After 24 h or 80 % confluence, cells were transfected with plasmid DNA (5 μg per well of a 6-well plate) using Lipofectamine 3000 (Thermo Fisher Scientific) at manufacturers specifications for 5 h, then replated into relevant culture plates.

### Live-Cell Fluorescent Microscopy

NSC34 cells expressing SOD1 constructs were cultured in 96-well plates in conditions previously mentioned for 72 h post transfection. Cells expressing SOD1-EGFP constructs were imaged at time points 24, 48 and 72 h post transfection on the incucyte live cell imager (Sartorius) with a 10 x objective and excitation wavelength 470 nm. Images were illumination corrected using CellProfiler^28^ software version 4.1 (Broad Institute, MIT Harvard) then analysed using CellPose^29^ cell segmentation algorithm to count GFP positive cells across timepoints similar to our previous work^18^.

### Quantification of Inclusions in NSC-34 cells

Transfected NSC-34 cells were fixed 48 h post-transfection. Plates were pre-fixed for 5 min via removal of half a volume of growth media and replacement with pre-warmed (37°C) 4% paraformaldehyde (PFA) in 1× PBS solution, resulting in a 2% PFA solution. Following this, the 2% PFA solution was completely removed and replaced with 4% PFA for 20 min for full fix. Residual PFA was then quenched using 100 mM Tris (pH 8) for 10 minutes while rocking. Cells were then permeabilized in 1× PBS 0.1 % Triton X-100 for 5 min followed by a 10 min incubation with 1:5000 Hoechst 33342 (Life Technologies) in 1× PBS 0.1 % Triton X-100 (TX-100) solution. Finally, cells were washed three times in 1× PBS before being immediately imaged or stored in the dark at 4 °C. All imaging was performed within 72 h of fixation.

Imaging was performed using a DMi8 epifluorescent microscope (Leica, Germany) fitted with a 10× air objective and software-based autofocus. 720 images were captured in total, with those that contained artefacts removed from analysis (less than 10 images). Images were analysed as described previously^18^.

### Fluorescence Correlation Spectroscopy

NSC-34 cells transfected with SOD1 variants were lysed 48 h after transfection using ice-cold 1× PBS 0.1% TX-100 with 1× protease inhibitors and centrifuged at 17,000 × *g* for 20 min at 4 °C to pellet insoluble material. Following this, the soluble fraction was pipetted onto clear 1.5# glass cover slips in an 8-well gasket chamber and sealed with a microscope slide. Samples for fluorescence correlation spectroscopy (FCS) were equilibrated to 25 °C in the microscope incubation chamber for at least 30 min prior to performing FCS. A FaLCon SP8 confocal microscope (Leica, Germany) with an incubation chamber (25 °C), pulsed white light laser, and 86× water immersion objective was used to perform FCS. The laser pulse rate was set to 80 MHz, laser power/transmission was 85%/20% for FCS experiments. We read a single point within a sample 3 times for 10 seconds, with 20 second gaps between reads.

### Limited Proteolysis using Proteinase K

NSC-34 cells were transfected as above and cell lysates were harvested in 1× PBS 0.1% TX-100 with 1× protease inhibitors 48 h post-transfection. 100 ng/ml of proteinase K was added to the cell lysates at room temperature to initiate proteolysis. Samples of the cell lysates were then harvested at 4 time points: 15, 30, 45 and 60 minutes after the initial addition of proteinase K. At each of these time points, phenylmethylsulfonyl fluoride was added at a final concentration of 5 mM to halt enzymatic activity. 4X SDS-PAGE reducing buffer was subsequently added to all samples preparing them for analysis via western blot.

### Immunoblotting of SOD1-EGFP from Cell Lysates

Cells were cultured in 6 well plates for 48 h post transfection then lysed in 1 % TX 50 mM Tris (pH 8) 1 % TX-100 with 1X protease inhibitor and 10 mg/ml N-Ethylmaleimide (NEM) (Sigma Aldrich) for disulphide status assays. Samples were then spun for 20 min at 17 000 g and supernatant was collected, flash frozen and stored at -80 C° until use. Samples were treated with reducing and non-reducing 4 x loading buffer then boiled for 5 min at 95 C° prior to being loaded onto stain-free TGX Any-kDa SDS-PAGE precast gels (Bio-Rad, Australia). Following electrophoresis, total protein on the gels was quantified using Criterion Stain Free Image (Bio-Rad, Australia) prior to transferring for immunoblotting. Gels were electrophoresed for 1 h at 160 V. Proteins were transferred onto methanol-activated Amersham Hybond 0.2 um polyvinylidene difluoride membranes (GE Healthcare, Australia) at 100 V for 1 h at 4 C°. Membranes were blocked in a blocking solution containing 5 % skim milk powder/Tris-buffered saline with 0.02 % Tween 20 (w/v) (TBST) for 1 h at room temperature. Subsequently membranes were incubated with polyclonal rabbit anti-GFP antibody (Abcam, ab290, 1:5000) in blocking solution overnight at 4 C°. Following primary antibody incubation, membranes were washed three times in TBST and then incubated with horseradish peroxidase-conjugated secondary antibody for anti-rabbit IgG (Dako, P0448; 1:5000) in blocking solution containing 5 % skim milk for 1 h at room temperature. To visualise bands, membranes were incubated with ECL (GE Healthcare, Australia) and imaged with an Amersham 600 RB imager (GE Healthcare, Australia) or (Other gel reader name).

Disulphide status, as a percentage, was calculated via the densitometry of the disulphide band over the total density of both bands combined using ImageJ version 1.54f^30^.

## Data Availability

All data produced in the present study are available upon reasonable request to the authors.

## Author Contributions

**Conceptualization:** LM, BT, AH; **Methodology:** LM, VS, MB, BT, AH; **Validation:** LM, VS, MB, BT, AH; **Formal analysis:** LM, VS, MB, BT, AH; **Investigation:** LM, VS, MB, BT, AH; **Resources:** LM, BT, AH; **Data curation:** LM, VS, MB, BT, AH; **Writing - original draft:** LM; **Writing - review & editing:** LM, VS, MB, BT, AH; **Visualization:** LM; **Supervision:** LM; **Project administration:** LM, BT, AH; **Funding acquisition:** LM.

## Acknowledgements

LM acknowledges funding from Motor Neuron Disease Australia and FightMND. The authors thank the patient and their family for their contribution to this work. The authors would like to pay deep respects to the late Professor Justin J. Yerbury (University of Wollongong, Australia), without whom this work would not have been done and authors LM, VS, and MB owe immeasurable gratitude.

